# Polymorphisms in the *ACE2* Locus Associate with Severity of COVID-19 Infection

**DOI:** 10.1101/2020.06.18.20135152

**Authors:** Luke Wooster, Christopher J. Nicholson, Haakon H. Sigurslid, Christian L. Lino Cardenas, Rajeev Malhotra

**Author notes:** **Corresponding author:** Rajeev Malhotra, MD, Massachusetts General Hospital, Yawkey 5700; 55 Fruit Street, Boston, MA 02114. Indicates co-first authors who contributed equally to the manuscript (Luke Wooster, Christopher J. Nicholson).

## Abstract

Data from clinical studies suggests a strong association between underlying cardiometabolic disease and worse outcomes in COVID-19. Given that the SARS-CoV-2 virus has a unique marked affinity to the human angiotensin-converting enzyme 2 (ACE2) receptor, one potential explanation behind this phenomenon may involve the higher expression of ACE2 receptor in these patients. Here, we analyzed association between polymorphisms in the ACE2 locus and COVID-19 severity in 62 patients found to be COVID-19 positive by polymerase chain reaction. Of these patients, 23 required hospitalization due to COVID-19 infection. Of 61 *ACE2* single nucleotide polymorphisms (SNPs) genotyped in this patient cohort, 10 were significantly associated with tissue expression of ACE2. Logistic regression adjusted for age and for sex identified six of these ten SNPs to be significantly associated with hospitalization. These results provide preliminary evidence of a genetic link between the *ACE2* genotype and COVID-19 disease severity and suggest that the *ACE2* genotype may inform COVID-19 risk stratification and need for more intense therapy.

## Introduction

The mechanisms linked to severity of coronavirus disease 2019 (COVID-19), which has thus far taken the lives of more than 400,000 people worldwide, are not yet understood. Data from clinical studies suggests a strong association between underlying cardiometabolic disease and worse outcomes in COVID-19.^1^ One potential explanation behind this phenomenon may involve the higher expression of angiotensin-converting enzyme 2 (ACE2) receptor in these patients.^2^ COVID-19 infection is caused by binding of the SARS-CoV-2 virus through its spike (S) proteins to the ACE2 receptor, principally in Type II alveolar cells of the lung.^3^ ACE2 is also highly expressed in cardiac tissue and vascular endothelial cells, which may therefore provide a mechanism by which SARS-CoV-2 promotes direct injury of these tissues.^4,5^ Since interactions between the virus and ACE2 receptor are critical for virus entry into host cells, genetic polymorphisms that regulate ACE2 expression and/or the strength of these interactions may affect COVID-19 severity.^6^ Recent studies aimed to compare the prevalence of *ACE2* functional coding variants in different populations with endemic COVID-19.^6-8^ However, to date there are no data showing a relationship between *ACE2* genetic variants and clinical outcomes with COVID-19. The present study aims to analyze novel associations between mRNA expression-altering genetic variants of human *ACE2* and clinical severity in patients with COVID-19.

## Methods and Findings

Consecutive patients diagnosed with COVID-19 throughout the Partners Hospital System who were previously genotyped as part of the Partners Biobank repository were studied. All genotyping was performed using the Illumina Infinium MEGA Consortium v1 SNP array. The Genotype-Tissue Expression (GTEx) project dataset (v8 release) was used to determine SNPs with a significant association with *ACE2* tissue mRNA expression (expression quantitative trait loci, eQTLs) with a minor allele frequency (MAF) of at least 5%.^9^ No coding variants were included in analysis as none met the threshold of 5% MAF in this cohort. *ACE2* is located on the X chromosome but has been shown to escape X inactivation.^10^ Therefore, logistic regression analysis adjusted for age and sex (Plink v1.07) was performed using allelic count where males were 0 or 1 and females 0, 1, or 2. We analyzed the effects of *ACE2* minor allele dosage on requirement for hospitalization due to COVID-19. This study was approved by the Partners institutional review board (IRB protocol #2020P000982).

Among 30,752 previously genotyped Caucasian patients in the Partners Biobank, 62 were found to be COVID-19 positive by polymerase chain reaction between 1^st^ March and 16^th^ May 2020. Hospitalization status was determined on 27^th^ May by review of the electronic medical records. The median (IQR) age of this cohort was 62 (52, 73) years and 44% were female (**Table 1**). Of these patients, 23 (37%) required hospitalization due to COVID-19 infection. Across this cohort, we performed a case/control genetic association analysis on *ACE2* polymorphisms.^9^ Of 61 *ACE2* single nucleotide polymorphisms (SNPs) genotyped in this patient cohort, 10 were significant eQTLs for *ACE2* in at least one tissue type (*P*<1E-4) with some exhibiting linkage disequilibrium (**Table 2**). Of the 10 SNPs, five polymorphisms (rs4240157, rs6632680, rs4830965, rs1476524, and rs2048683), with minor alleles that associated with higher tissue expression of *ACE2*,^9^ were associated with increased need for hospitalization due to COVID-19 even after adjusting for age and sex (odds ratio 2.9-4.3, *P*<0.05 for all). Similarly, an *ACE2* polymorphism (rs1548474) whose minor allele associated with lower tissue expression was associated with lesser COVID-19 severity not requiring hospitalization (odds ratio 0.27, 95% CI 0.08-0.85, *P*=0.025, **Table 3**).

**Table 1.** Demographic and clinical characteristics among Caucasian patients who did or did not require hospitalization due to COVID-19.

| Characteristic | Total | Required Hospitalization |  | P value |
| --- | --- | --- | --- | --- |
|  |  | No | Yes |  |
| No. | 62 | 39 | 23 |  |
| Age, median (IQR) | 62 (52, 73) | 61 (49, 68) | 71 (59, 74) | 0.061 |
| Female, n (%) | 27 (43.5) | 18 (46.2) | 9 (39.1) | 0.609 |
| Diabetes, n (%) | 26 (41.9) | 15 (38.5) | 11 (47.8) | 0.596 |
| CVD, n (%) | 21 (33.9) | 11 (28.2) | 10 (43.5) | 0.271 |
| Hyperlipidemia, n (%) | 43 (69.4) | 26 (66.7) | 17 (73.9) | 0.584 |
| Hypertension*, n (%) | 47 (75.8) | 26 (66.7) | 21 (91.3) | 0.034 |
| CKD, n (%) | 10 (16.1) | 7 (17.9) | 3 (13.0) | 0.731 |

CVD = cardiovascular disease; CKD = chronic kidney disease
For age, the Mann Whitney test was used. For the remaining categorical variables, Fisher's exact test was used.
\* The presence of hypertension was not associated with any of the 10 SNPs in Table 2 by Fisher's exact test.

**Table 2.**
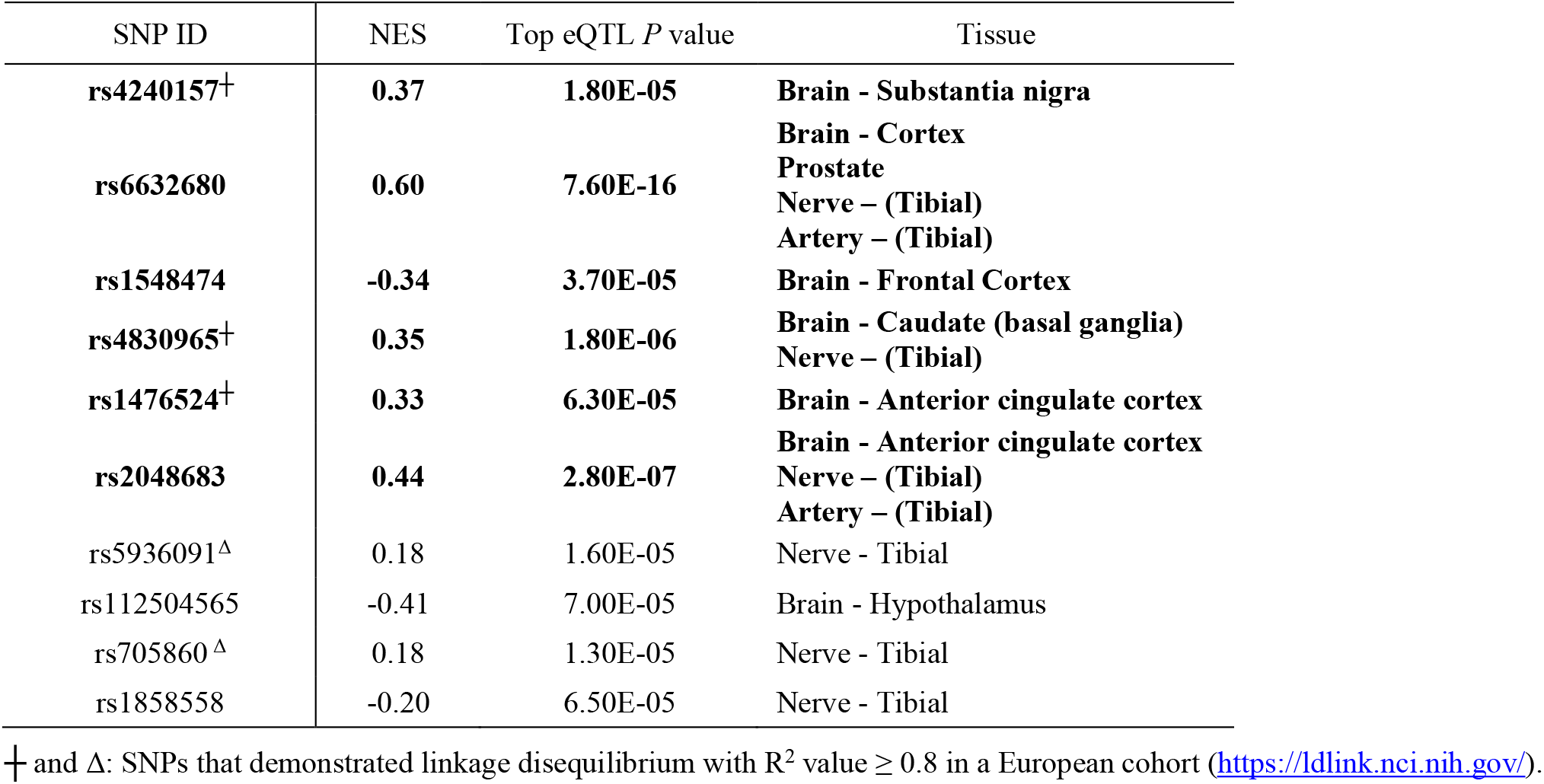
Association of *ACE2* eQTLs with ACE2 Tissue Expression.

**Table 3.** Association of *ACE2* eQTLs with need for hospitalization in COVID-19 patients.

| SNP ID | Major Allele | Minor Allele | MAF in Hospitalized | MAF in Non-Hospitalized | Multivariable Logistic Regression OR* | OR 95% Confidence Interval | Multivariable Logistic Regression *P* value* |
| --- | --- | --- | --- | --- | --- | --- | --- |
| **rs4240157<sup>+</sup>** | **C** | **T** | **0.56** | **0.33** | **4.28** | **(1.34, 13.69)** | **0.014** |
| **rs6632680** | **A** | **C** | **0.62** | **0.31** | **4.05** | **(1.32, 12.37)** | **0.014** |
| **rs1548474** | **G** | **T** | **0.19** | **0.42** | **0.27** | **(0.08, 0.85)** | **0.025** |
| **rs4830965<sup>+</sup>** | **A** | **G** | **0.50** | **0.28** | **3.45** | **(1.15, 10.29)** | **0.027** |
| **rs1476524<sup>+</sup>** | **T** | **C** | **0.50** | **0.30** | **3.41** | **(1.12, 10.28)** | **0.030** |
| **rs2048683** | **T** | **G** | **0.50** | **0.30** | **2.94** | **(1.02, 8.55)** | **0.047** |
| rs5936091<sup>Δ</sup> | G | A | 0.28 | 0.17 | 1.63 | (0.56, 4.73) | 0.371 |
| rs112504565 | C | T | 0.14 | 0.17 | 0.85 | (0.24, 2.95) | 0.797 |
| rs705860<sup>Δ</sup> | A | G | 0.19 | 0.16 | 1.14 | (0.37, 3.53) | 0.819 |
| rs1858558 | T | A | 0.13 | 0.11 | 1.09 | (0.26, 4.59) | 0.910 |

MAF = minor allele frequency; OR = odds ratio for requiring hospitalization; NES = Normalized effect size
\*adjusted for Age and Sex
<sup>+</sup> and <sup>Δ</sup>: SNPs that demonstrated linkage disequilibrium with $R^2$ value $\geq 0.8$ in a European cohort (https://ldlink.nci.nih.gov/).

## Discussion

Previous studies have investigated the potential for ACE2 polymorphisms to explain population-based differences in COVID-19 severity. Cao et al. found no direct evidence of population differences in coronavirus S-protein binding-resistant mutants. However, considerably higher allele frequencies for SNPs associated with higher *ACE2* expression in tissues were observed in the East Asian populations.^6^ Asselta et al. reported no significant differences in the burden of rare deleterious *ACE2* variants when comparing an Italian population with Europeans and East Asians.^7^ In contrast, using single cell sequencing, Zhao et al. demonstrated that *ACE2* was more abundantly expressed in type II alveolar cells from an Asian male donor, when compared to white and African American donors.^8^ Whilst these studies shed light on the contribution of *ACE2* genetic variation on population-based differences in coronavirus etiology, our analysis is the first to identify *ACE2* polymorphisms that may influence disease severity. Importantly, genetic variants that have been associated with *ACE2* mRNA expression levels associated with disease severity. Further studies that analyze the expression altering effects of these polymorphisms in lung and cardiovascular tissue will be informative to fully understand their role in COVID-19 infectivity and disease severity.

## Conclusion

In summary, in a cohort of patients with COVID-19 and pre-existing genotypic data, we found that six out of ten eQTL variants in *ACE2*, that are associated with altered *ACE2* tissue expression, were associated with the need for hospitalization due to COVID-19. These results, although requiring replication in a validation cohort and in non-Caucasian populations, provide preliminary evidence of a genetic link between the *ACE2* genotype and COVID-19 disease severity and suggest that the *ACE2* genotype may inform COVID-19 risk stratification and need for more intense therapy.

## Data Availability

All data referred to in this manuscript will be made available upon request to the full extent allowable by Partners HealthCare policy.

https://www.gtexportal.org/home/

## Acknowledgements

The authors have no conflicts of interest to disclose. Dr. Malhotra is supported by NHLBI R01 HL142809, American Heart Association grant 18TPA34230025, and the Wild Family Foundation.

